# Impact of Exercise-induced Dehydration on Perceived Exertion During Endurance Exercise: A Systematic Review with Meta-analysis

**DOI:** 10.1101/2021.11.12.21266279

**Authors:** Thomas A. Deshayes, Timothée Pancrate, Eric D.B. Goulet

**Author notes:** Correspondence: Eric D.B. Goulet, Ph.D. Performance, Hydration and Thermoregulation Laboratory University of Sherbrooke 2500 boul. de l’Université Sherbrooke, Québec, Canada J1K 2R1 *62728.

## Abstract

Understanding the impact of stressors on the rating of perceived exertion (RPE) is relevant from a performance and exercise adherence/participation standpoint. Athletes and recreationally active individuals dehydrate during exercise. No attempt has been made to systematically determine the impact of exercise-induced dehydration (EID) on RPE. The present meta-analysis aimed to determine the effect of EID on RPE during endurance exercise and examine the moderating effect of potential confounders using a meta-analytical approach. Data analyses were performed on raw RPE values using random-effects models weighted mean effect summaries and meta-regressions with robust standard errors, and with a practical meaningful effect set at 1 point difference between euhydration (EUH) and EID. Only controlled crossover studies measuring RPE with a Borg scale in healthy adults performing ≥ 30 min of continuous endurance exercise while dehydrating or drinking to maintain EUH were included. Sixteen studies were included, representing 147 individuals. Mean body mass loss with EUH was 0.5 ± 0.4%, compared to 2.3 ± 0.5% with EID (range 1.7 to 3.1%). Within an EID of 0.5 to 3% body mass, a maximum difference in RPE of 0.81 points (95% CI: 0.36-1.27) was observed between conditions. A meta-regression revealed that RPE increases by 0.21 points for each 1% increase in EID (95% CI: 0.12-0.31). Humidity, ambient temperature and aerobic capacity did not alter the relationship between EID and RPE. Therefore, the effect of EID on RPE is unlikely to be practically meaningful until a body mass loss of at least 3%.

## INTRODUCTION

Rating of perceived exertion (RPE), a subjective estimation of the intensity or difficulty of a physical task, is widely used by professionals in the field of exercise sciences, coaching and sports medicine to monitor or prescribe exercise intensity (87). Developed by Gunnar Borg (19, 20), the Borg RPE scale is a universally accessible, comprehensible, useful, non-invasive, valid and inexpensive tool that can be used in diverse populations such as in children, adolescents, young and older adults, and under different conditions, including leisure and elite sports, clinical rehabilitation and scientific research. Although the etiology of RPE is unclear, it is proposed that it is either centrally derived (64) or generated by neuronal processes that integrate afferent signals from various peripheral and central sources, as well as from psychological factors (20, 88).

The RPE is a pivotal component of aerobic exercise. Indeed, it acts as a regulator of exercise intensity (1, 93) and exercise duration (26, 35, 77) and, thus, modulates exercise capacity in competitive athletes and, as important, is at the core of the decision to engage and adhere to the regular practice of physical activity among recreationally active individuals (32, 33). Given that physiological and psychological signals can act individually or in concert to disturb RPE, it follows that limiting the number of physiological or emotional stressors to a minimum during exercise should ensure optimal performance for the athlete and lead to a sense of fulfillment in recreationally active individuals (1).

Depending on a host of factors, sweat losses typically reach 0.5 to 1.7 L/h during exercise (13). Athletes as well as recreationally active individuals do not usually replace all their fluid losses during exercise. Exercise-induced dehydration (EID), best represented by the acute body mass loss accrued during exercise, alters thermoregulatory, metabolic and cardiovascular functions (24, 43, 62, 63, 71), more particularly in individuals with low aerobic fitness (69) and may predispose to the development of thirst, headaches, tiredness, mental fatigue (44) and impaired mood (6, 39), while its impact upon cognitive performance is still debated (45). These factors may contribute to increasing RPE during exercise, ultimately impeding exercise performance in athletes (55) and potentially decreasing exercise adherence and participation in recreationally active individuals (33, 82) which, from a societal and health perspective, is not suitable. Indeed, both the affective response and RPE are associated with long-term physical activity participation (97).

The relationship between EID and RPE has received much scientific attention, with some studies showing that EID can significantly increase RPE (2, 14, 16, 37, 38, 42, 43, 54, 56, 62, 63, 71, 72, 75, 95, 99), while others did not (8, 12, 17, 28, 30, 31, 34, 40, 51, 53, 59, 61, 68, 73, 74, 84, 89). At this time, no attempt has been made to systematically determine the impact of EID on RPE. Discrepancies between findings could potentially be related to methodological differences among studies, albeit this remains to be determined and confirmed with the aid of relevant analyses.

Efforts have yet to be deployed to determine, using a meta-analytic approach, the impact of EID on RPE. More specifically, there is a need to answer these questions: 1) does the change in RPE during exercise relate to EID?; 2) what is the magnitude of the effect of EID on RPE across different levels of EID; 3) is the magnitude of the effect of EID on RPE practically important?; 4) are there any identifiable factors among ambient temperature, humidity level, exercise intensity, exercise duration and aerobic capacity that may moderate the relationship between EID and RPE and; 5) to which extent “cardiovascular strain” mediates the relationship between EID and RPE? The goal of this study, therefore, is to use a meta-analytic approach to provide answers to the above-mentioned questions. Findings will be valuable to scientists, physical trainers, sports nutritionists, physicians, exercise physiologists, sports psychologists and any individuals engaged in regular exercise. Also, such findings will be valuable to elucidate whether the changes in body water are part of an integrated signal.

## METHODS

### Experimental approach to the problem

Although the relationship between EID and RPE has received scientific attention, results are controversial. No attempt has been made to summarize and determine the magnitude of changes in RPE across different levels of EID. Such a gap indicates a need to use a systematic approach with meta-analysis to further our understanding of the relationship between these variables.

Figure 1 reports the search strategy used for study selection. The literature search, limited to original peer-reviewed articles published in French or English, was performed with the PubMed, MEDLINE, SPORTDiscus, AMED and CINAHL databases, combining a “title field” and an “abstract field” research using the following keywords alone or in combination: cycling, dehydration, drink, effort, endurance, euhydration, exercise, exertion, fluid, hydration, hypohydration, perceived effort, perceived exertion, perception, performance, rate of perceived exertion, rating of perceived exertion, RPE and running. The exact search strategy can be found in supplementary material 1. A first selection based on the title was performed; afterward, the abstract and method sections of all potential articles were read. When hydration status was manipulated and RPE measured, the methodological section was carefully read to verify eligibility. Published abstracts, case studies, non-peer-review manuscripts and conference proceedings were not considered. Cross-referencing was performed on included studies and 6 narrative/systematic reviews (3, 23, 48, 55, 66, 79). When needed, authors of included studies were contacted and asked to share experimental raw data. The last search of the literature was done on December 17, 2020. The review and the protocol were not registered. The meta-analysis was conducted using the Preferred Reporting Items for Systematic Reviews and Meta-analysis (PRISMA) guidelines.

**Figure 1.**
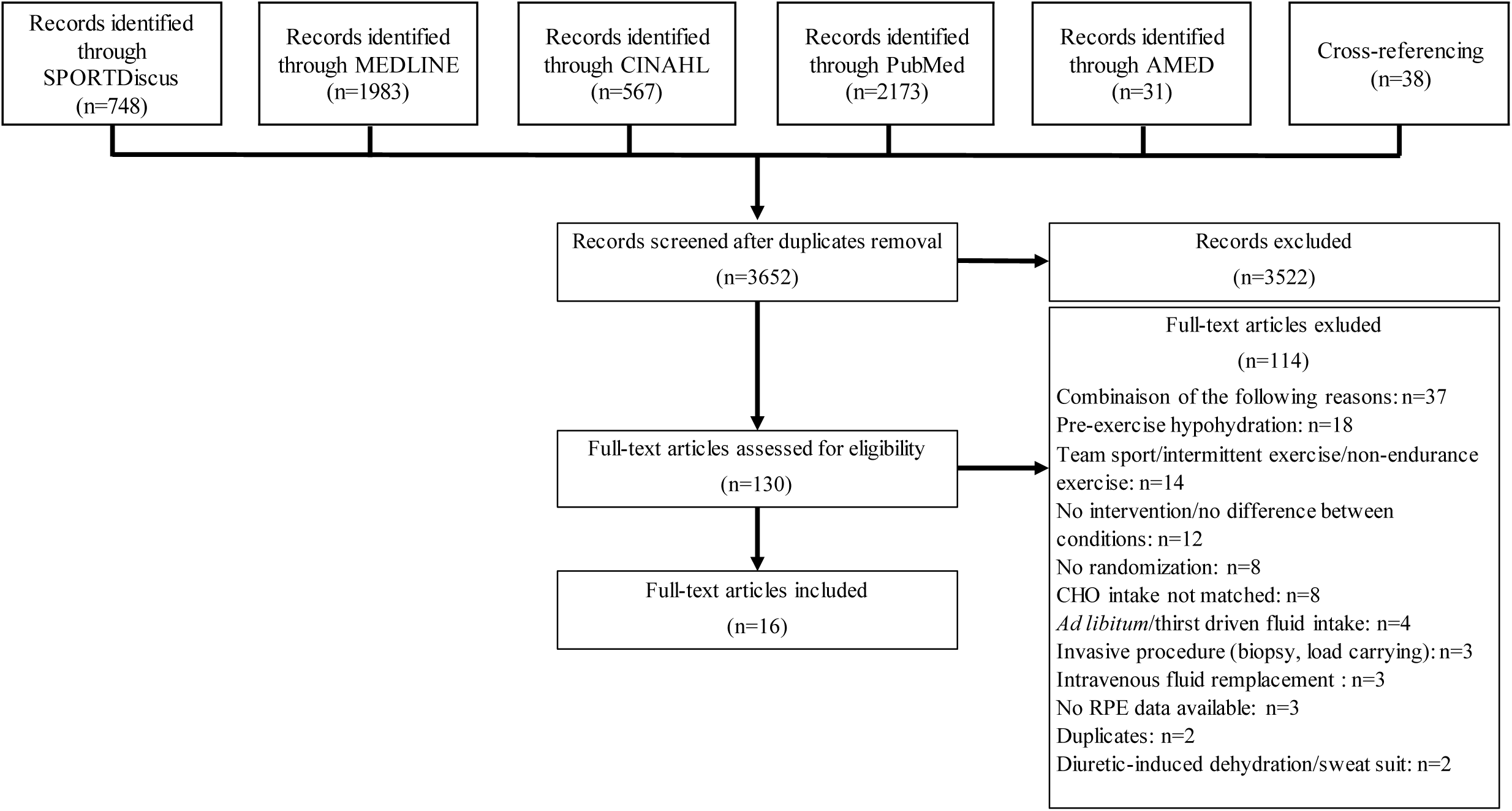
Flowchart showing the selection process used for the inclusion and exclusion of studies.

### Criteria for considering studies for inclusion in the meta-analysis

Inclusion criteria were: (1) controlled study using a randomized crossover design in healthy adults (≥ 18 years old); (2) ≥ 30 min of continuous running or cycling endurance exercise; (3) final EID in the experimental group > 1% of pre-exercise body mass and ≥ 0.5% than the euhydrated (EUH) control condition; (4) dehydration progressively induced during, not before exercise (46); (5) body mass change with EUH was within -1 to + 0.5% of the pre-exercise body mass (46); (6) fluid replacement was given orally; (7) if carbohydrates (CHO) or caffeine were provided during exercise, the amount was identical between conditions (9, 29) and; (8) data required to compute changes in EID and RPE included. Exclusion criteria were: (1) sports-specific and intermittent exercises; (2) use of diuretics or sweatsuit to accelerate EID; (3) provision of fluid according to thirst sensation; (4) uncontrolled ambient conditions or experimentation timing; (5) collection of muscle biopsies and; (6) carrying of loads during exercise.

### Assessment of trial quality

No specific and validated tool to assess the quality of exercise-related studies has been developed. Moreover, assessing trial quality in meta-analyses using a scale can influence the interpretation of results (57). Hence, trial quality assessment was not performed in the present meta-analysis.

### Data extraction

Using double data entry, data regarding (1) study characteristics; (2) participants characteristics; (3) exercise protocol characteristics; (4) EID levels and; (5) RPE were extracted and coded in spreadsheets. When not provided by authors, data only available in figures were extracted using WebPlotDigitizer.

### Exercise duration and intensity and participants’ ⩒O_2max_

Exercise duration was computed as the average exercise time completed during both the EID and EUH conditions. Exercise intensity was taken as the average of the mean % ⩒O_2max_ at which both the EID and EUH conditions were performed. Mean exercise intensity was computed using a weighted average for those studies that used a combination of exercise intensities. When not measured by authors, exercise intensity was estimated and computed as explained by Goulet (46). Most studies (4, 8, 16, 25, 31, 38, 68, 74, 75, 84, 89, 94, 95, 98) reported participants’ ⩒O_2max_; Barwood, Goodall and Bateman (15) and Dugas, Oosthuizen, Tucker, et al. (30) did not, and these values were calculated as in Goulet (46).

### Fluid intake, exercise-induced dehydration and dehydration rate measurement

Hydration rate (mL/min) was computed as the total amount of fluid intake divided by exercise duration, with the relative hydration rate (mL/min/kg) corrected for pre-exercise body mass (kg). The percent change in body mass from the pre- to post-exercise period was used as an index of the level of dehydration incurred during exercise. While this index is an imperfect representation of EID (65) as it is impacted by both metabolic water production and gas exchange during exercise, measurement error is relatively low amounting to an overestimation of fluid loss of ∼ 100 mL/h during moderate intensity exercise.

When not provided, pre-exercise body mass was taken as that provided in the sample description, whereas % body mass loss was computed using the following equation:

Pre-exercise body mass (kg) - post-exercise body mass (kg) / pre-exercise body mass (kg) x 100 (1)

Thus, any positive value represents a body mass loss while negative values indicate body mass gain.

Assuming a high repeatability of (11), and consistency in (60), sweating rate and thus body mass loss (22) during exercise at a given intensity, dehydration rate (% body mass loss/min) was computed as follows:

End of exercise body mass loss (%) / exercise duration (min) (2)

Dehydration rate was used to calculate the % body mass loss associated with each measurement of RPE within a single study. For example, if in a study RPE was measured at 30, 60 and 90 min and the dehydration rate was 0.03%/min, therefore the corresponding % body mass losses were respectively taken as 0.9 (ex., 30 min x 0.03%/min), 1.8 and 2.7%. In some studies, this iteration process had to be stopped when body mass loss surpassed 1% in the EUH condition (8, 31, 38, 74, 94). This procedure enabled us to pinpoint the behavior of RPE across a wide range of % body mass loss changes, using research data available from all included studies. To provide a practical, easy to understand and clear visual characterization of the effect of body mass loss on RPE during exercise, Figures 2a and b present the relationship between % body mass loss and RPE at fixed and anchored body mass loss levels, according to the classification presented in Table 1.

**Figure 2.**
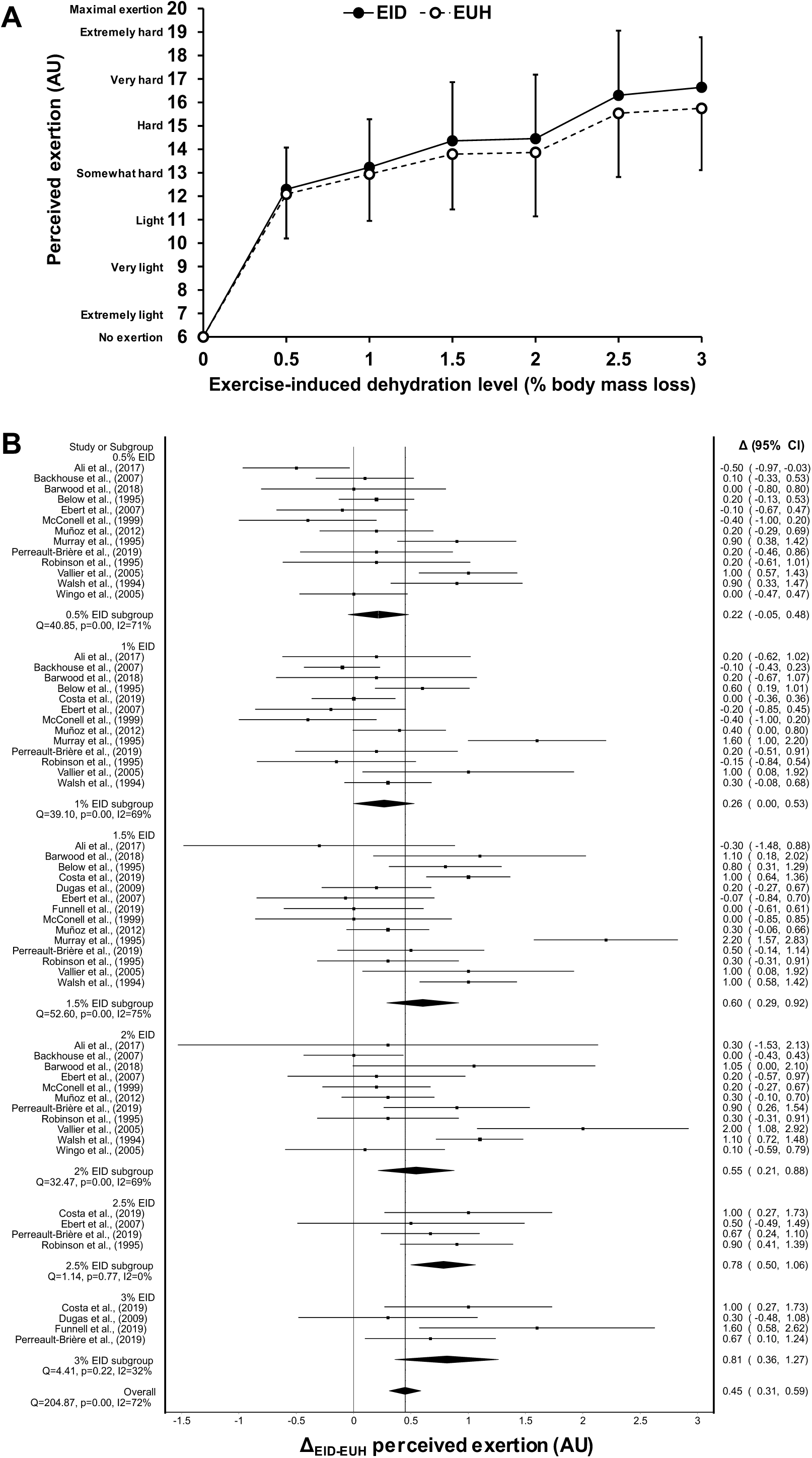
(a) Changes in perceived exertion (means ± SD) occurring during exercise between the control (EUH) and exercise-induced dehydration (EID) condition across levels of exercise-induced dehydration of 0.5, 1, 1.5, 2, 2.5 and 3% body mass. AU: arbitrary units. EID: exercise-induced dehydration. EUH: euhydration. (b) Forest plot showing the mean differences in perceived exertion across different levels of exercise-induced dehydration. Filled diamond symbol represents the weighted mean change in perceived exertion between conditions. Size of squares is proportional to the weight of each study. CI: confidence interval.

**Table 1.** Classification of percent body mass losses.

| Classification |  |  |
| --- | --- | --- |
| From | to | corresponds to |
| 0.26 | 0.75% | 0.5% |
| 0.76 | 1.25% | 1.0% |
| 1.26 | 1.75% | 1.5% |
| 1.76 | 2.25% | 2.0% |
| 2.26 | 2.75% | 2.5% |
| 2.76 | 3.25% | 3.0% |

### Measurement of perceived exertion during exercise

Perceived exertion data are presented according to the original 6-20 Borg scale. Dugas, Oosthuizen, Tucker, et al. (30) and Walsh, Noakes, Hawley, et al. (95) used the Borg- CR10 scale (range 0-10); for those studies, RPE was converted back to the 6-20 scale according to Arney, Glover, Fusco, et al. (7). For Barwood, Goodall and Bateman (15) (hot and cold drink conditions), Dugas, Oosthuizen, Tucker, et al. (30) (0, 33 and 66% conditions) and Murray, Michael and McClellan (75) (5- and 10-min conditions) merging of experiments were performed to eliminate data dependency.

### Moderating variables

The following variables were *a priori* identified as potential moderators for the relationship between RPE and EID: ambient temperature, relative humidity, exercise duration and intensity and participants’ ⩒O_2max_. Ambient temperature and relative humidity are interdependent, as are exercise intensity and duration. To take this into account, absolute humidity and a composite score of exercise stress (product of exercise intensity (% ⩒O_2max_) and exercise duration (min)) were also considered as moderating variables.

### Mediating variable

Heart rate was considered a potential mediating variable regarding the relationship between EID and RPE. Core temperature would have been another one to consider, but the paucity of data prevented us from evaluating its impact. For each study, mean exercise heart rate difference between the EUH and EID conditions was computed by averaging the sum of the heart rate difference computed at each measurement point.

### Statistical analyses

#### Software

Data were analyzed in their original form using Microsoft Office Excel 2020 (version 1902, Redmond, WA, USA), MetaXL, Comprehensive Meta-Analysis (version 2.2.064, Englewood, NJ, USA), STATA/MP (version 14, College Station, TX, USA), SPSS macros provided by Lipsey and Watson (46, 47) and IBM SPSS Statistics (version 21, Armonk, NY, USA) software.

#### Weighted mean effect summaries

Each of the studies included in the meta-analysis took measurements of RPE during exercise at more than one EID level. Therefore, a meta-analysis of repeated measures was performed in an effort to limit the violation of the assumption of data independence in the data structure (85). First, an all-points forest plot was constructed to determine the mean effect of EID on RPE at body mass loss levels fixed and anchored at 0.5, 1, 1.5, 2, 2.5 and 3%. This method allows us to illustrate the rate of increase in RPE across this range of body mass losses. Moreover, it allows for more precision in establishing the relationship between RPE and body mass loss as this strategy increases the n for any dehydration point. All RPE data within a given EID level were independent of each other; however, each study contributed in providing RPE-related data to more than one EID level. Nevertheless, the assumption of independence was protected for each of the EID levels. Post-hoc analyses were done using the False Discovery Rate procedure, with the number of *a priori* defined comparisons taken as 6, mirroring each of the EID levels compared. Second, to establish the mean effect of EID on RPE for each increase in 1% body mass loss, a forest plot was constructed from the slope estimates of the relationships between EID and RPE for each of the included studies, using non-weighted linear regression analyses with the intercepts forced through the origin (85). For this forest plot, *n* was taken as the number of EID levels included in the regression analysis. Initially, the analyses were performed separately by subgroups. However, data from studies using time-trial type exercise protocols were combined with those using fixed-intensity exercise protocols given the low number of studies using time-trial type exercise and because variations in RPE within the different EID levels were similar and, in all instances, < 1 point. Nonetheless, on few occasions, analyses excluding time-trial type exercises will be presented when deemed interesting. Weighted mean effect summaries were determined using method of moment random-effects model. A more intuitive approach was used to verify whether it would change the outcomes in comparison to our approach. For that, we averaged, within a given hydration condition, all RPE measurements across time, and then observed the difference in RPE between conditions.

#### Practical significance of the weighted mean effect summaries and slope estimate

The qualitative interpretation of the practical significance of the effect of EID on RPE was performed as in Goulet & Hoffman (48). Previous studies observed reliability of the RPE scale to be < 1 point (41, 58). Because the minimal increment of the scale is 1 point, this threshold was taken and accepted as the smallest worthwhile practical difference in RPE.

#### Heterogeneity, publication bias and sensitivity analysis

Cochran’s Q and I² statistic were both used to assess between-study heterogeneity and the degree of inconsistency among results of included studies. Cochran’s Q test was considered significant at *p* ≤ 0.1 (18). The following classification was used to interpret the I² statistic: low (< 40%), moderate (40-59%), substantial (> 60%) (52). Publication bias was performed using visual assessment of funnel plots with Trim and Fill adjustments. A sensitivity analysis was performed on each of the forest plots by removing each study once from the models to determine whether this would change the magnitude of the outcome summaries.

#### Meta-regression analyses

The potential mediating effect of heart rate and moderating effect of the *a priori* defined confounders were determined by regressing the slope estimates upon the mean heart rate difference between conditions or each of the confounders, respectively. Confounder and mediator variables included at least 10 data points from 10 different studies. A multiple meta-regression combining all moderators (with the exception of absolute humidity and composite score of exercise stress) was performed to understand the strength of our proposed model. Meta-regression analyses were performed using method of moment random-effects model, with 95% robust (Huber-Eicher-White-sandwich) standard errors.

#### Statistical significance

Otherwise stated, in all instances, results were considered significant at *p* < 0.05 or when the 95% confidence interval did not include 0.

#### Variance computations

When raw data were obtained from the authors, variances were directly calculated from the Δ standard errors or standard deviations of the absolute changes in RPE between conditions. Otherwise, individual variances for changes in RPE were estimated as in Goulet and Hoffman (48) using an imputed weighted correlation coefficient of 0.81 deriving from 40 correlation coefficients obtained from 5 different studies whose authors provided raw data.

## RESULTS

### Search results and characteristics of the included studies

After removing duplicates, 3652 titles were checked (Figure 1). In the remaining 130 articles assessed for eligibility, 16 were included in the meta-analysis (Table 2). The studies were published between 1994 and 2019 in 10 different peer-reviewed journals. Four studies were conducted in the USA (16, 74, 75, 98), 3 in the UK (8, 15, 38), Australia (25, 31, 68), and South-Africa (30, 89, 95) and 1 in Canada (84), France (94) and New-Zealand (4).

**Table 2.**
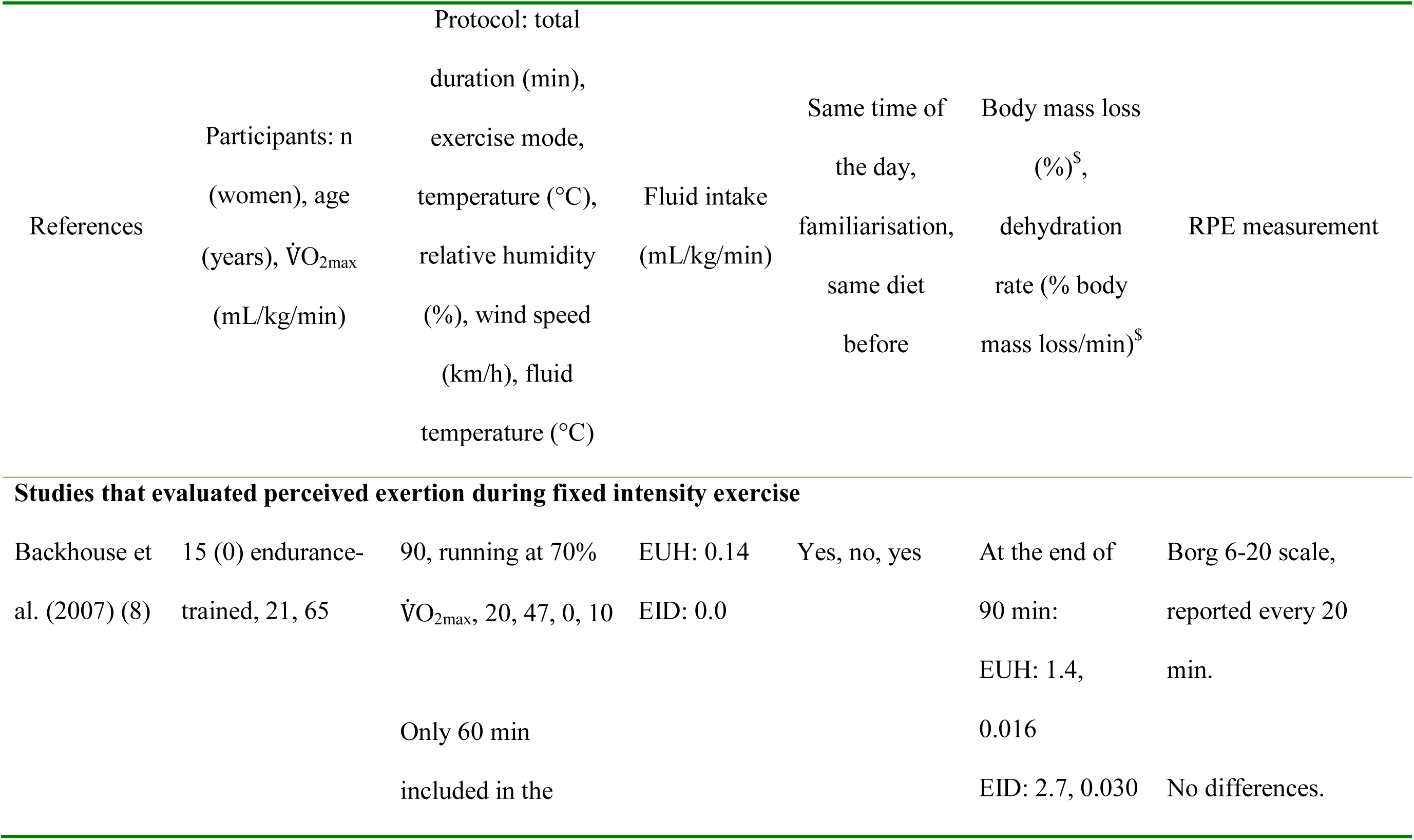
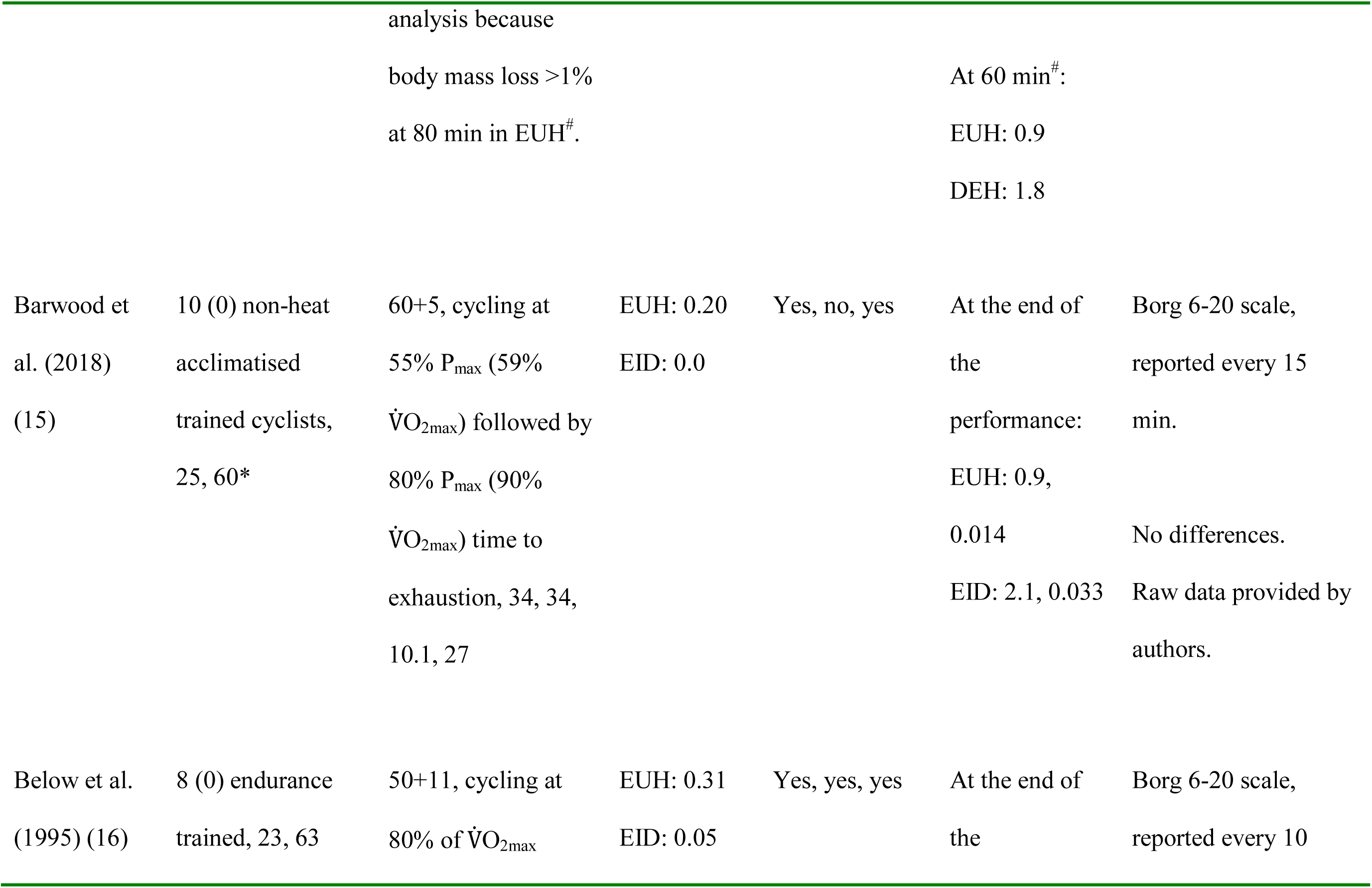
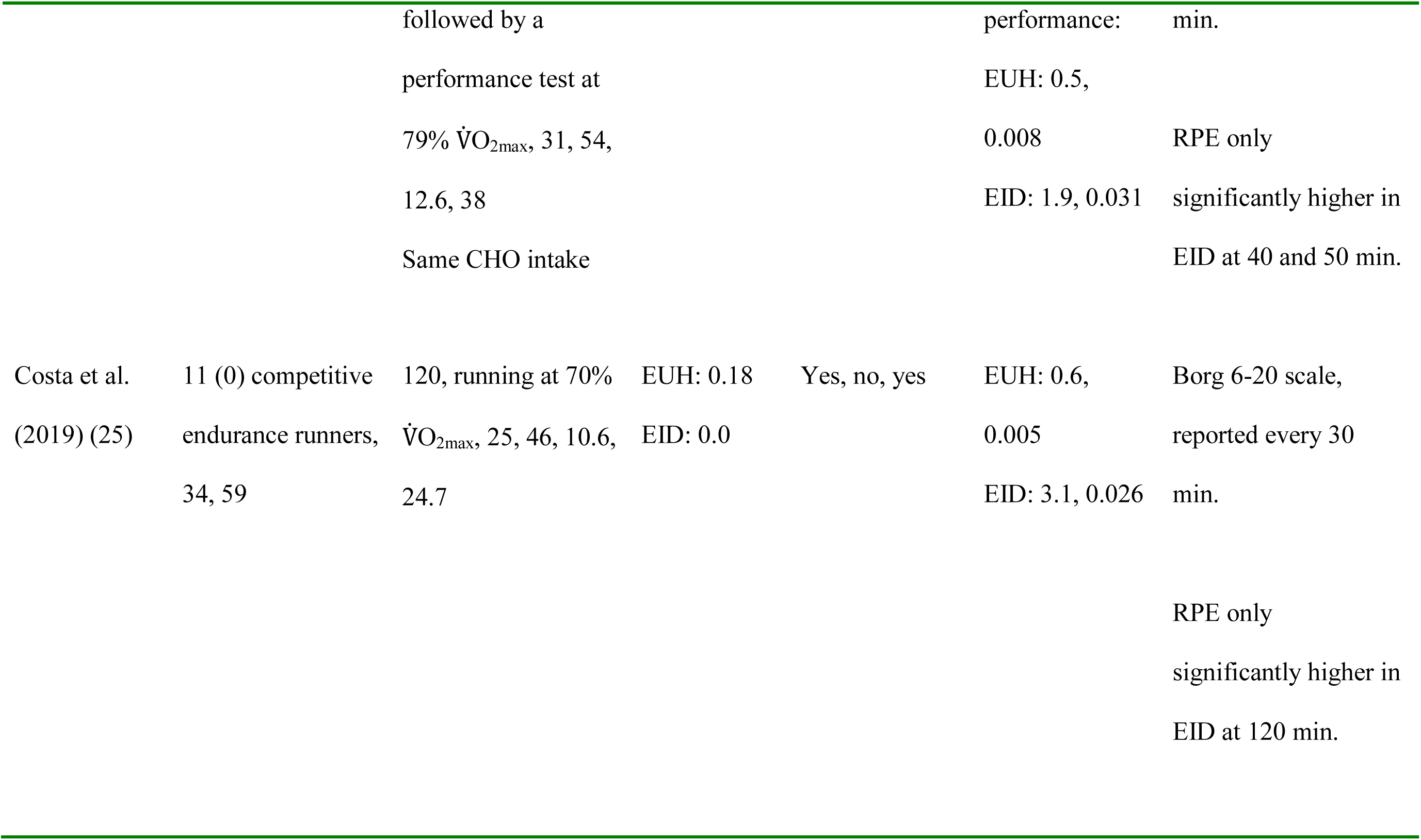
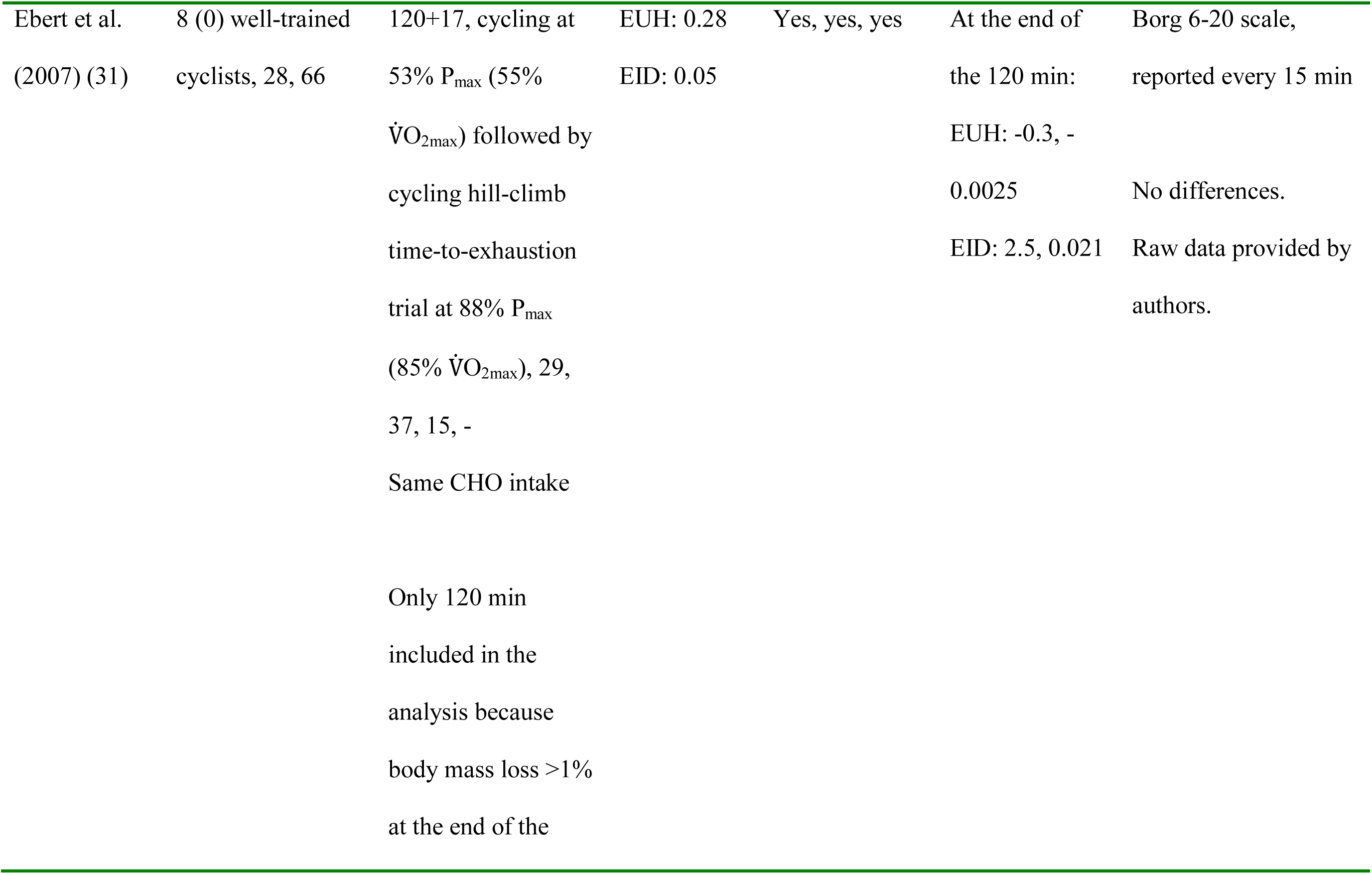
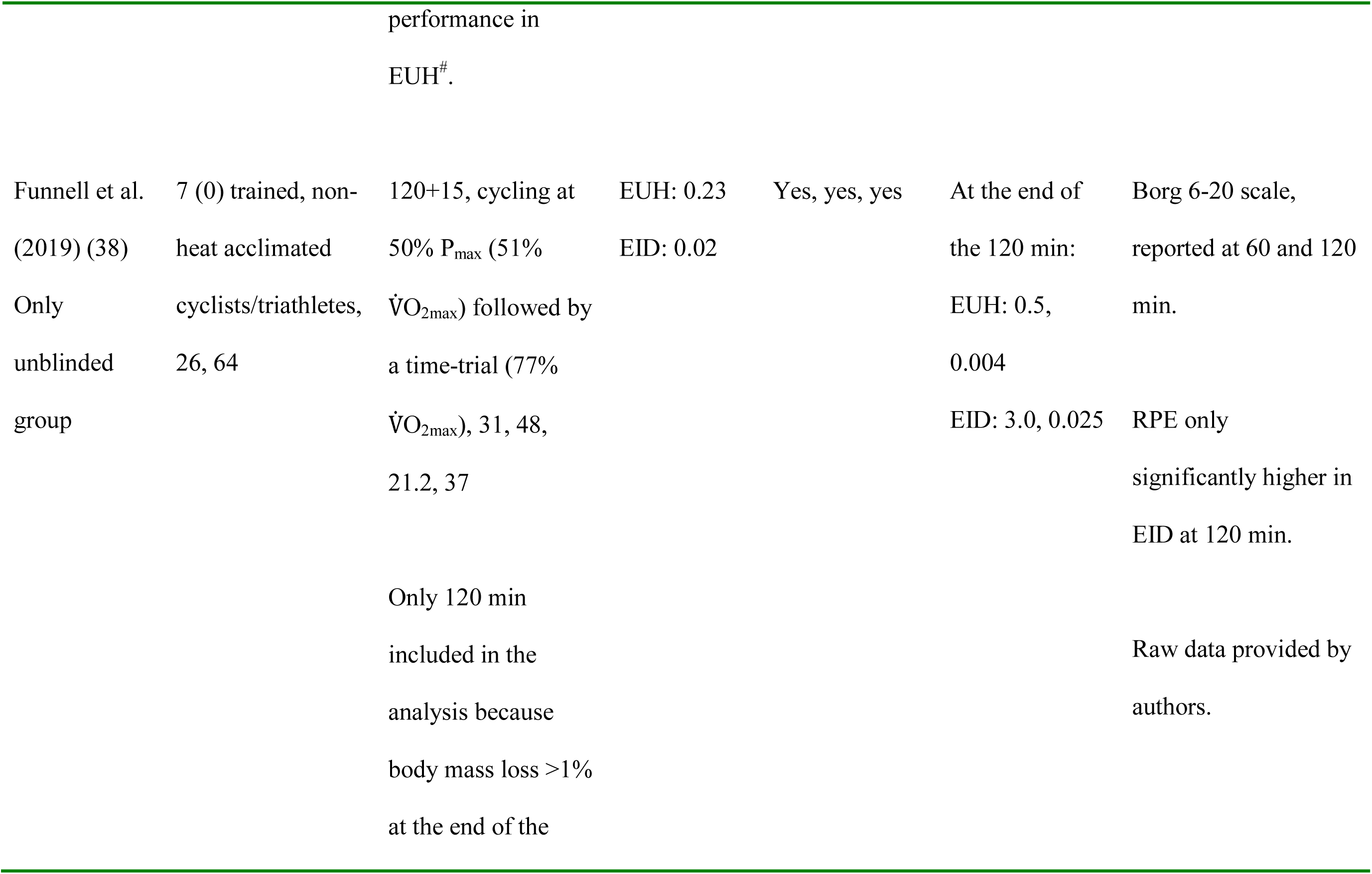
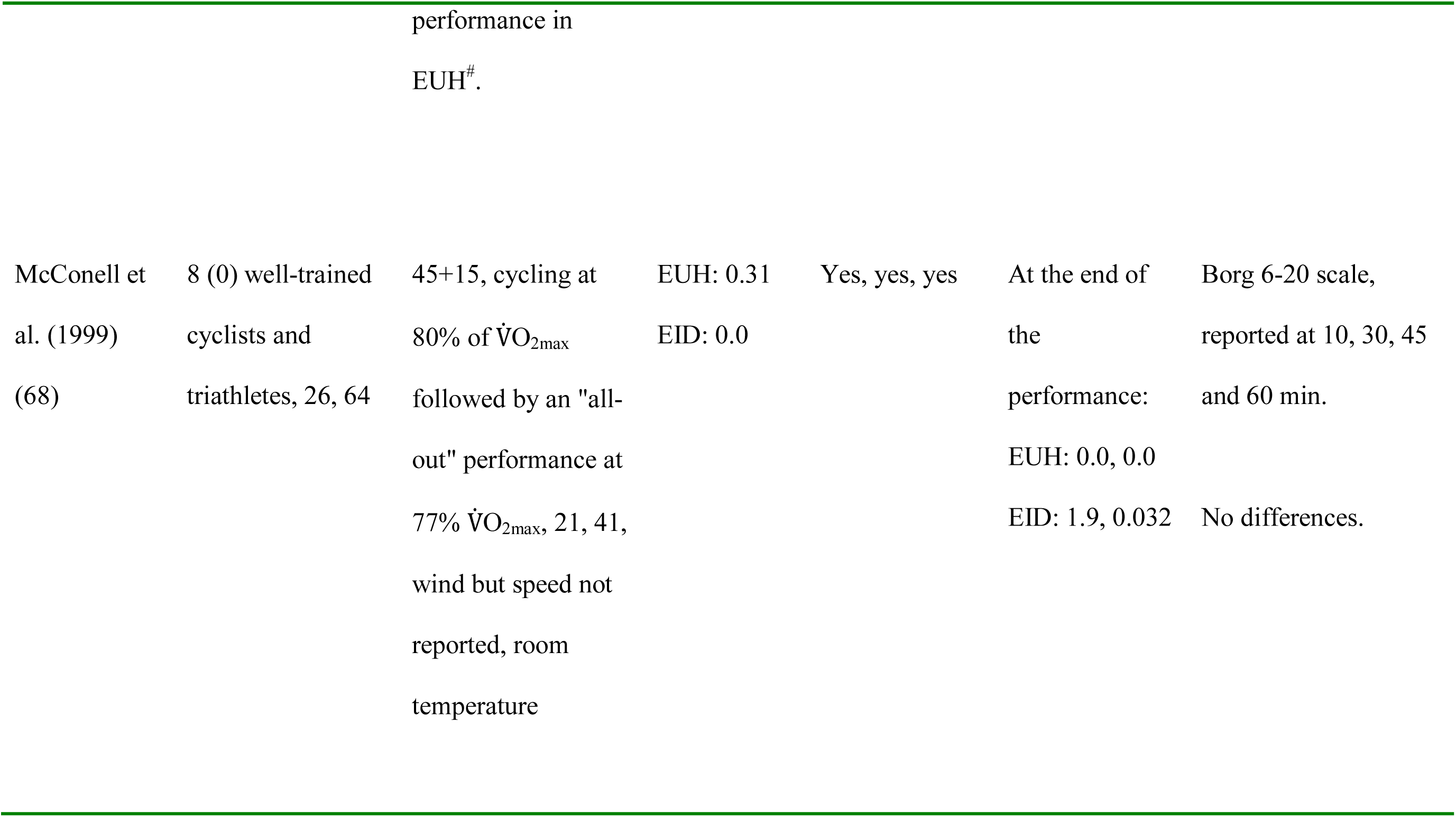
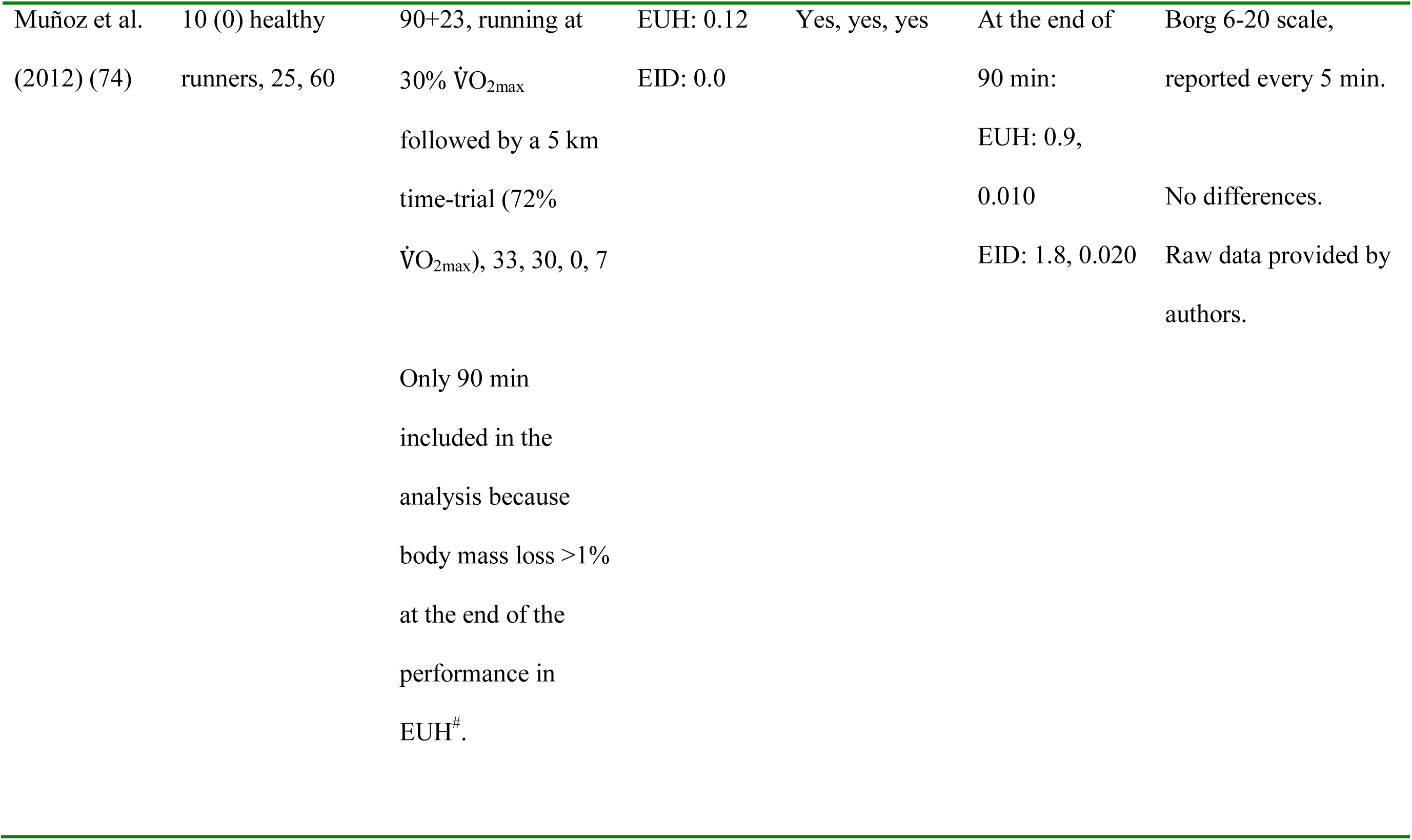
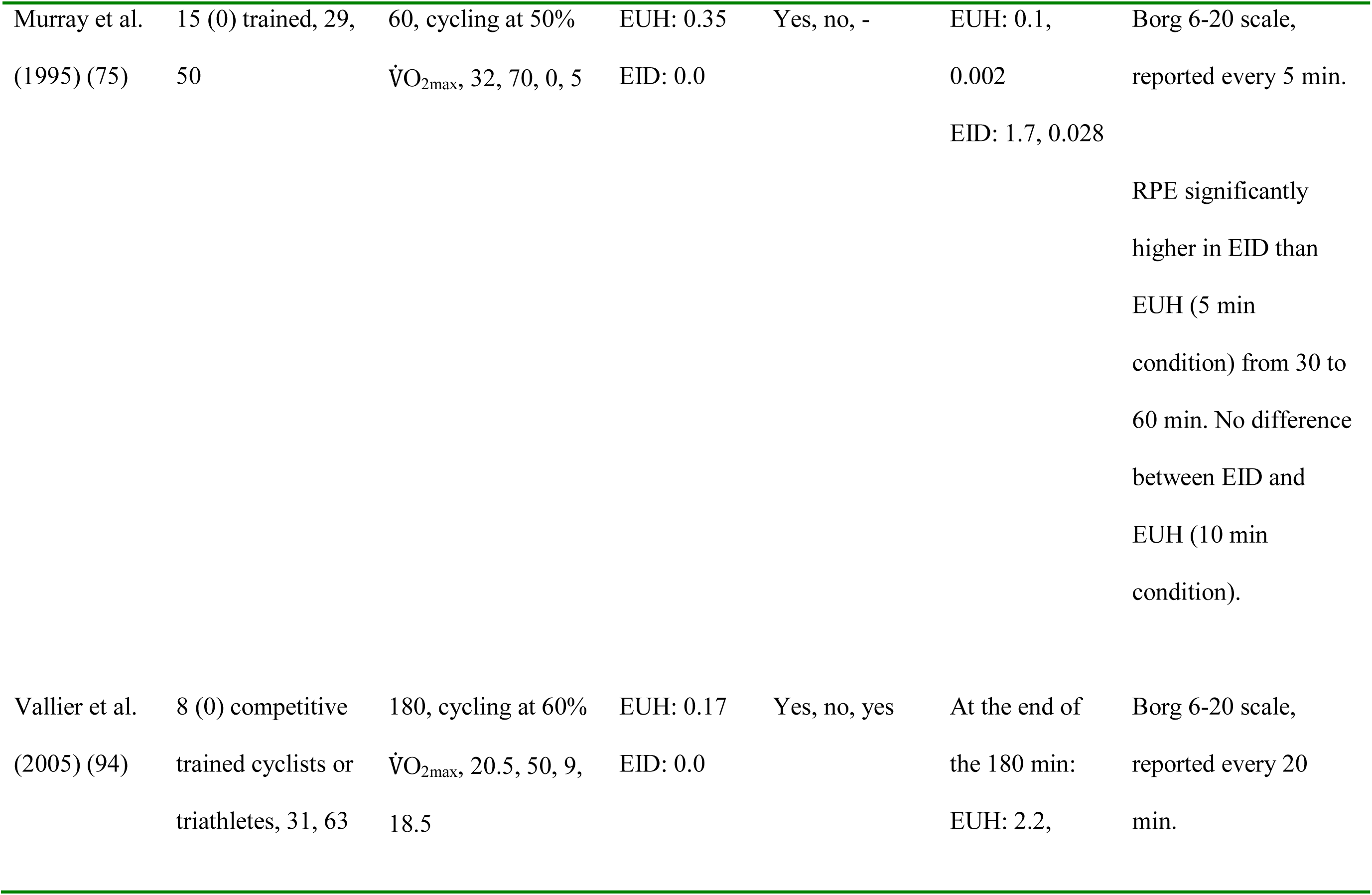
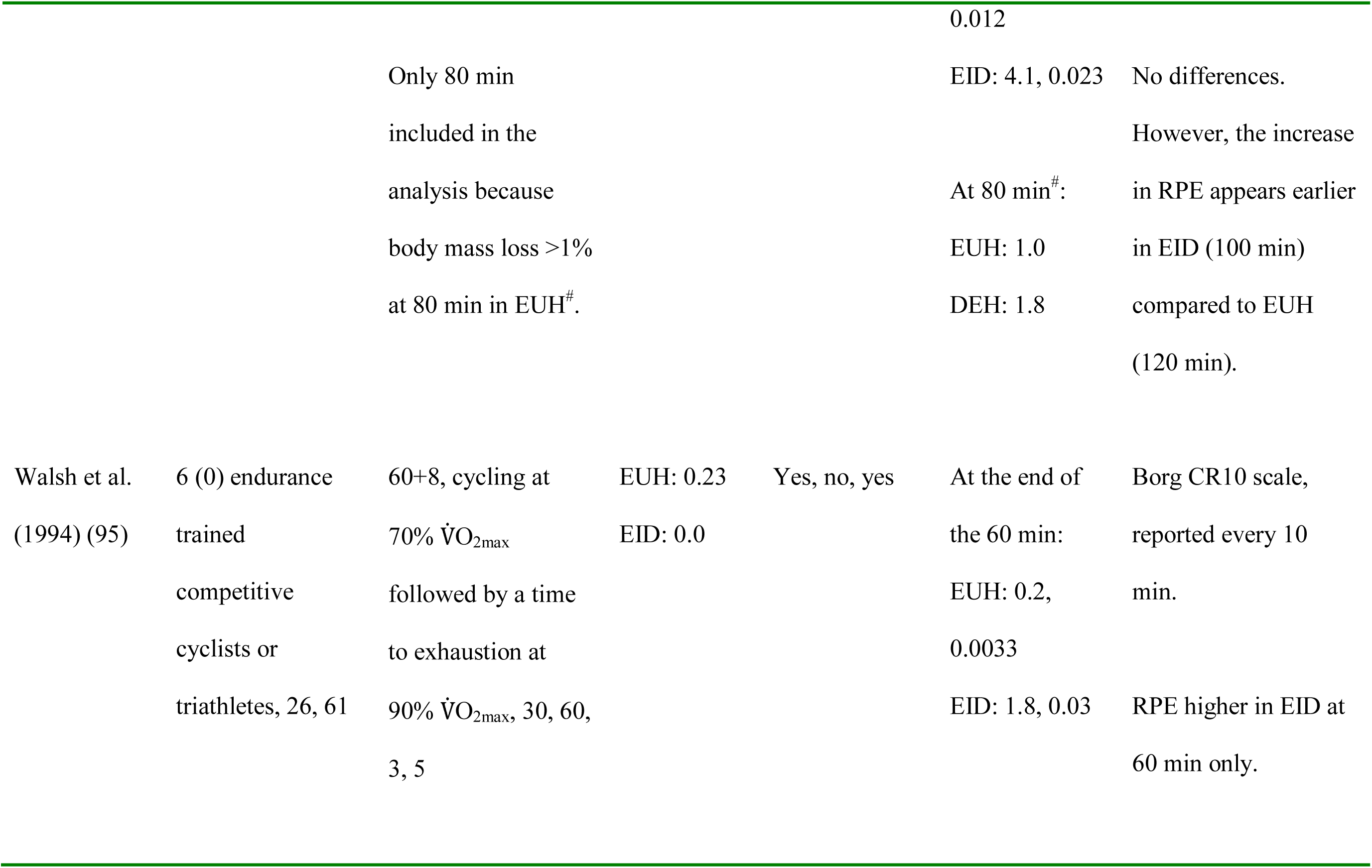
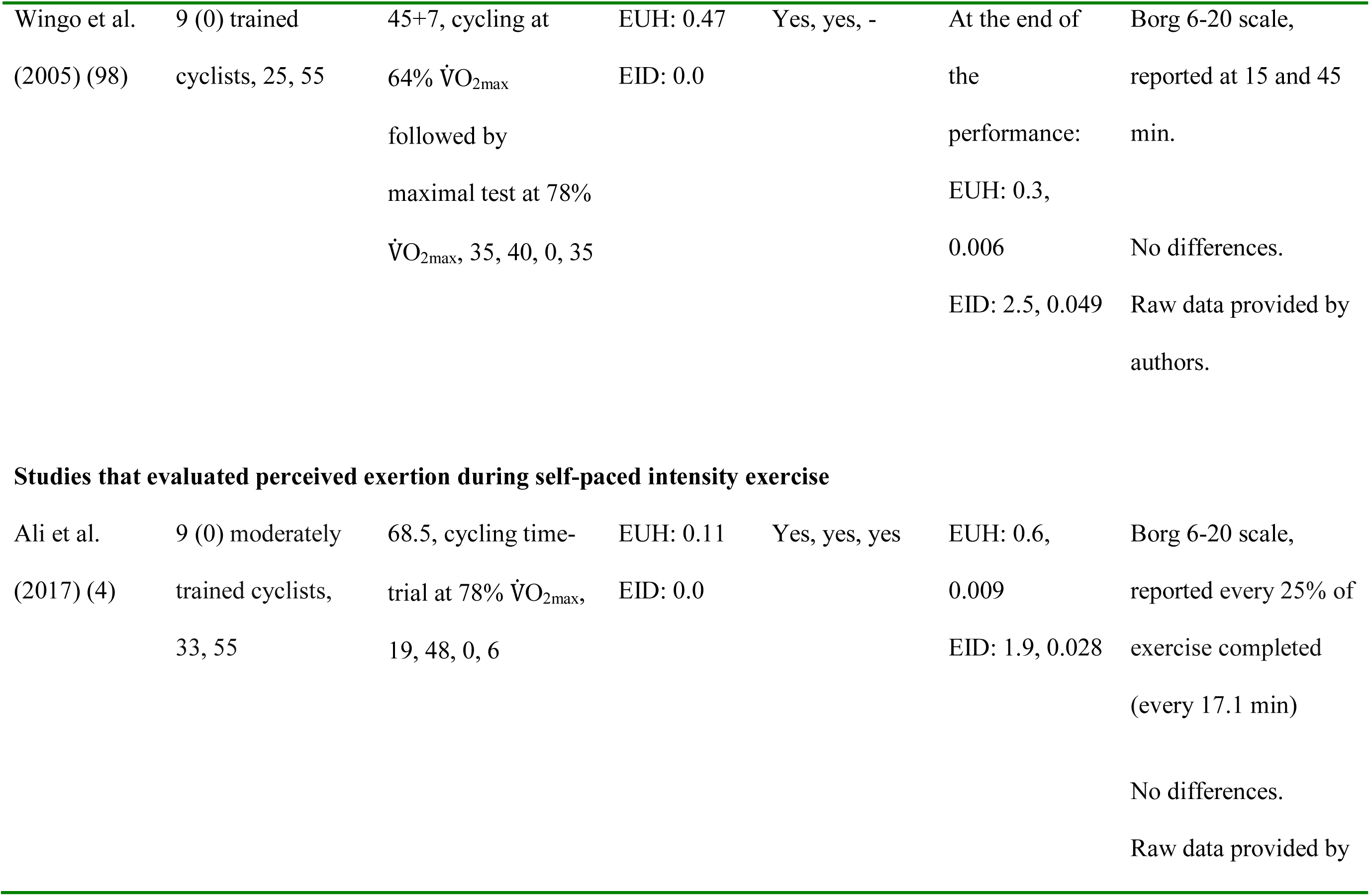
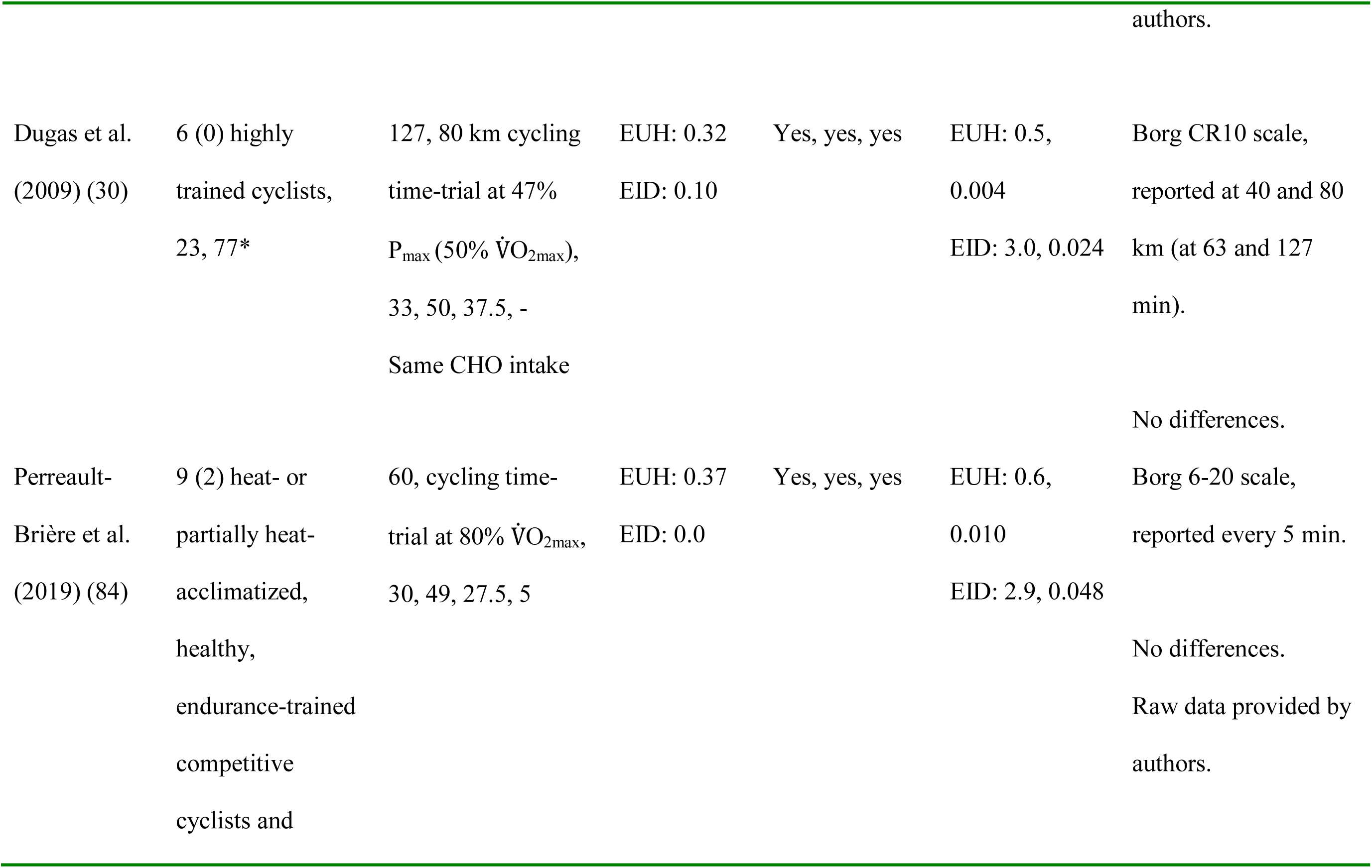
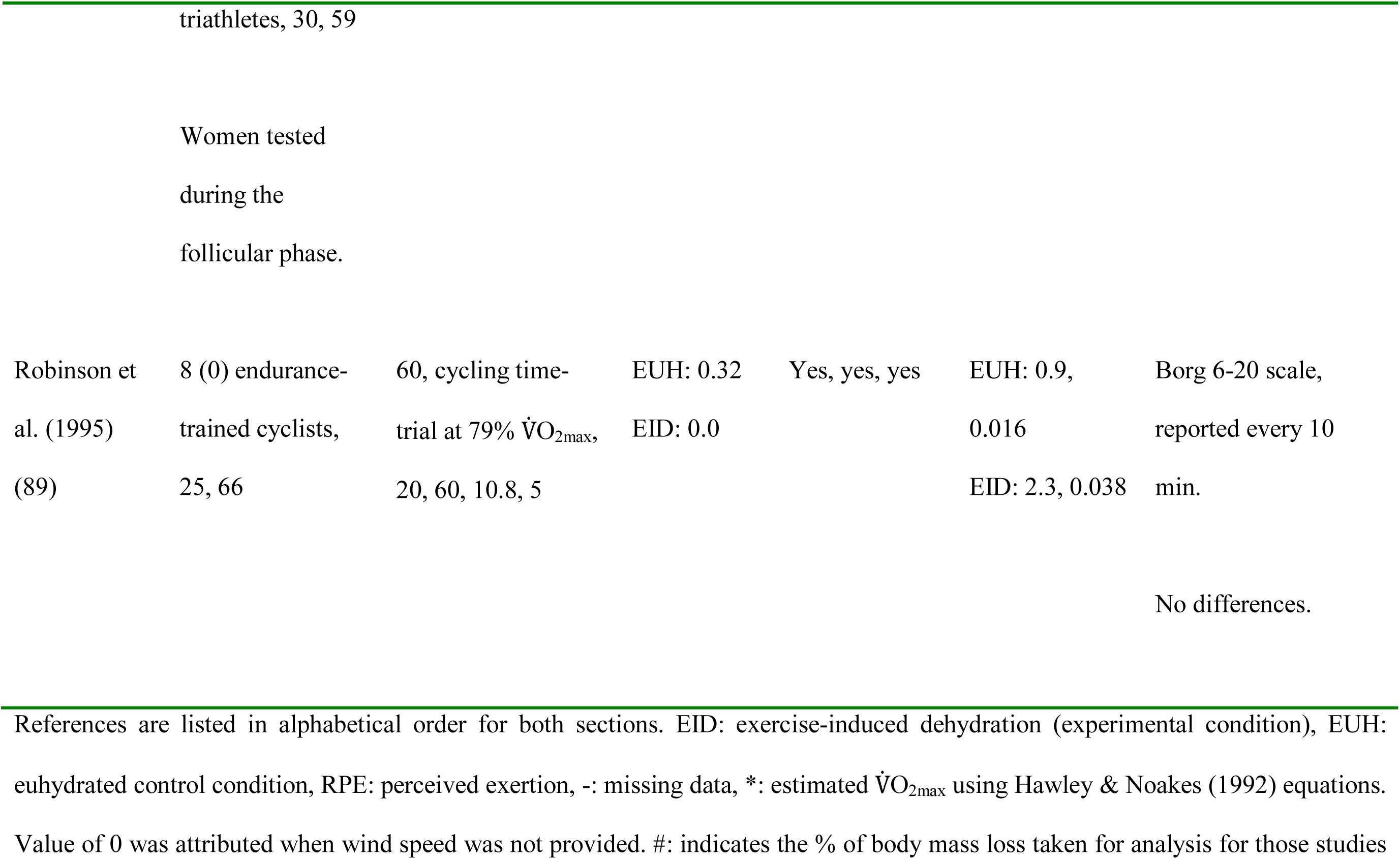
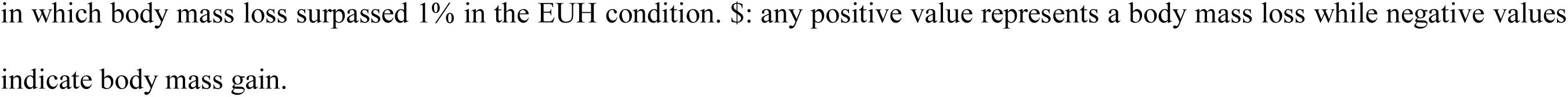
Summary of characteristics of included studies.

### Participant’s characteristics

A total of 147 endurance-trained individuals are represented among the 16 included studies, with women representing only 1% of the total sample. Mean sample size was 9 ± 3 individuals per study (range 6-15). None of the included studies reported information about ethnicity. The mean age, height, body mass, body mass index, ⩒O_2max_ and peak power output of the participants were respectively 27 ± 4 years, 179 ± 2 cm, 73 ± 3 kg, 23 ± 1 kg/m^2^, 62 ± 6 mL/kg/min and 389 ± 39 W.

### Characteristics of the exercise protocols

Among the selected studies, 81% (n = 13) used cycling as the mode of exercise while the remaining used running (n = 3). The mean ambient temperature and relative humidity were respectively 28 ± 6°C and 48 ± 10%, with a mean wind speed of 10 ± 11 km/h. The mean exercise duration and intensity were respectively 79 ± 27 min (range 51-127 min) and 65 ± 13% of ⩒O_2max_.

### Fluid consumption and exercise-induced dehydration levels

Mean rates of fluid consumption in the EUH and EID conditions were respectively 18.9 ± 7.5 and 1.0 ± 2.0 mL/min, representing 0.26 ± 0.1 and 0.01 ± 0.03 mL/kg/min. The average fluid temperature was 17 ± 13°C. The mean end-of-exercise body mass loss was 0.5 ± 0.4% (range 1 to -0.3%) when EUH was attempted to be maintained, compared to 2.3 ± 0.5% (range 3.1 to 1.7%) with EID, for a mean difference of 1.7 ± 0.7% (range 2.8 to 0.9%) between conditions. Mean dehydration rates of 0.007 ± 0.005 and 0.03 ± 0.009%/min were observed during the EUH and EID conditions, respectively.

### Weighted mean effect summaries

Figure 2a depicts the changes in RPE that occurred during exercise between the EUH and EID conditions across levels of body mass losses of 0.5, 1, 1.5, 2, 2.5 and 3%, and for absolute values of RPE which fluctuated from ∼ 12 (light) to 16.5 (hard) points. Figure 2b pinpoints the weighted mean difference in RPE between the EUH and EID conditions across each of these levels of body mass losses. Results of the forest plot illustrate that, compared with EUH, EID slowly increased RPE during exercise from 0.22 points (95% CI: -0.05-0.48) when body mass loss was trivial (0.5%) to 0.60 points (95% CI: 0.29- 0.92) when body mass loss was light (1.5%), up to 0.81 points (95% CI: 0.36-1.27) when body mass loss was moderate (3%). Only at 0.5 and 1% body mass losses were the differences in RPE between the EUH and EID conditions not significant. In none of the 6 weighted mean summary effects models did the removal of each study one at a time significantly and practically impacted the outcome that body mass loss has upon RPE. For each of the EID subgroups, the practical impact of body mass loss on RPE was likely or almost certainly trivial. Distribution of point estimates around each of the 6 weighted mean effect summaries was appropriate, which indicates no publication bias. Cluster analysis indicates that heterogeneity was substantial with an *I*^2^ of 72% and a Cochran’s *Q* of 204.9, *p* < 0.01. At the subgroup level, substantial inconsistencies were also observed at the 0.5, 1, 1.5 and 2, but not 2.5 and 3% body mass loss levels where heterogeneity was low. When the analyses were performed without the studies that used time-trials, results were similar, with the exception that the differences in RPE between the EUH and EID condition reached 1.2 points (95% CI: 0.61-1.80) when body mass loss was moderate (3%).

Cluster analysis indicates that EID, on average, increases RPE by 0.45 point (95% CI: 0.31-0.59, Figure 2b). Using the more intuitive approach, we observed an overall effect of 0.38 points (95% CI: 0.22-0.53, Q = 125.3, *p* < 0.01, I^2^ = 88%). When studies that used time-trials were removed, the results were, again, extremely similar: 0.50 points (95% CI: 0.33-0.67, Q = 174.8, *p* < 0.01, I^2^ = 77%) *vs.* 0.44 points (95% CI: 0.24-0.64, Q = 114.2, *p* < 0.01, I^2^ = 90%) with the more intuitive approach.

Figure 3a illustrates the slope estimates for the regression of RPE on the % body mass loss for each of the included studies. Figure 3b shows a forest plot combining the 16 slope estimates to derive a weighted mean summary effect. Results show that for each 1% body mass loss, RPE increased on average by 0.21 points (95% CI: 0.12-0.31), thereby theoretically suggesting that it is not before reaching a body mass loss of ∼ 5% that EID may potentially affect RPE in a meaningful way. A sensitivity analysis revealed that the removal of each study one at a time from the model did not significantly nor practically alter the outcome of the weighted mean effect summary, with variations in the slope estimate ranging from 0.13 (95% CI: 0.06-0.20) to 0.35 points (95% CI: 0.17-0.53). Inconsistency among research observations was substantial with an *I*^2^ of 75% and a Cochran’s *Q* of 58.99, *p* < 0.01. Point estimates were not equally distributed on each side of the weighted mean summary effect, thereby suggesting publication bias. A trim and fill analysis adjusting for missing studies at the left side of the mean changed the weighted mean effect summary to 0.10 points (95% CI: 0.00-0.22). When the analyses were performed without the studies that used time-trials, results showed that for each 1% body mass loss, RPE increased on average by 0.38 points (95% CI: 0.17-0.59, Q = 56.2, *p* < 0.01, I^2^ = 80%). Figure 3c depicts the relationship between RPE and % body mass loss while including all 59 study-specific data points, which violates the assumption of independence among data. Nevertheless, the weighted regression analysis provides a slope estimate (0.26 points, 95% CI: 0.10-0.42) which is congruent to the one built from combining all 16 slope estimates.

**Figure 3.**
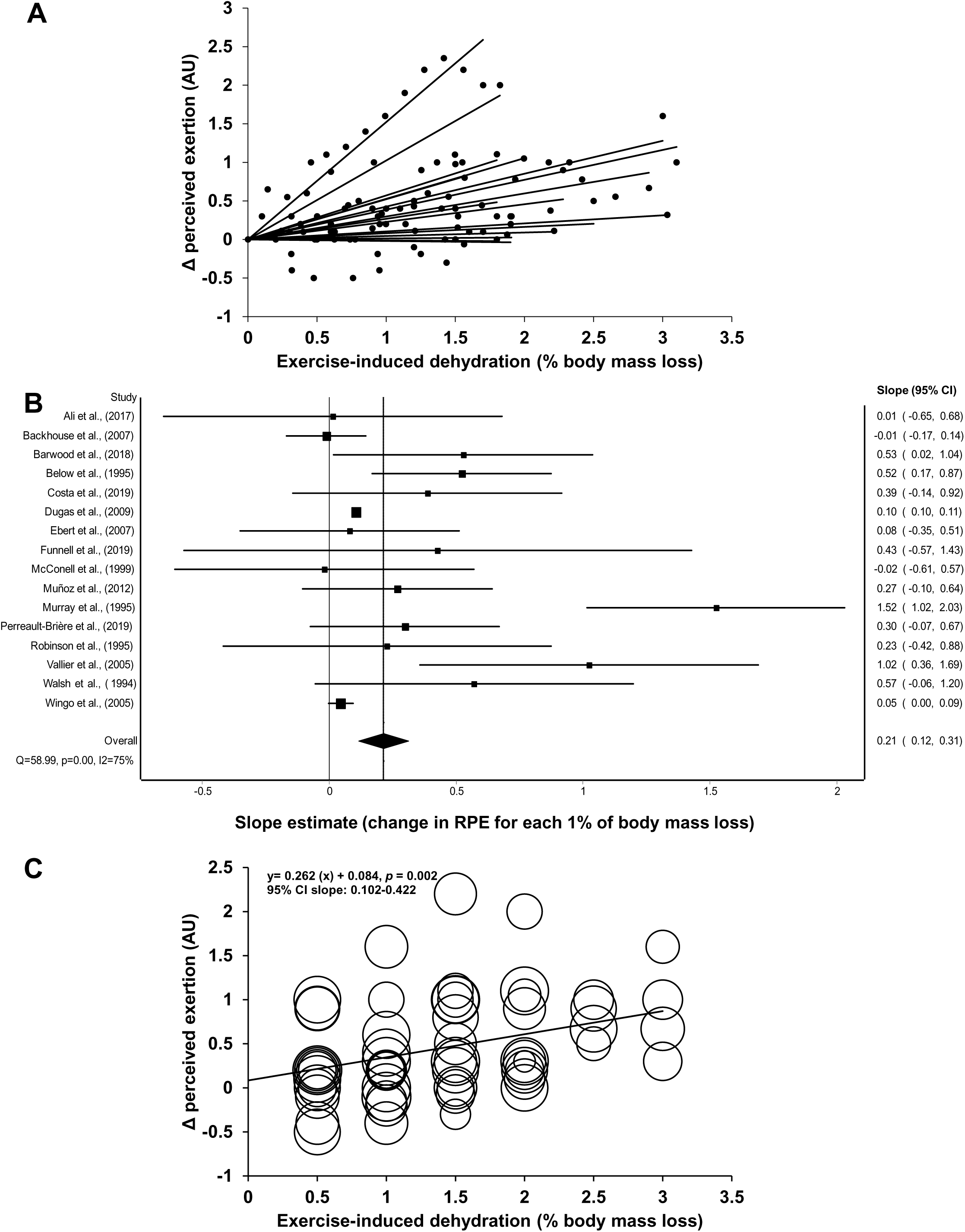
(a) Slope estimates for the regression of perceived exertion on the % body mass loss for each of the included studies; (b) Forest plot combining all slope estimates to derive a weighted mean summary effect; (c) Relationship between perceived exertion and % body mass loss while including all study-specific data points.

### Meta-regression analyses

Figure 4 shows the relationships between the changes in slope estimates and (a) temperature, (b) humidity level, (c) exercise duration, (d) exercise intensity, (e) aerobic capacity and (f) mean heart rate difference across the different studies. Individually, none of these variables significantly correlated to the extent of changes in RPE for each 1% in body mass loss. When all these variables were combined in a multiple meta-regression model (except heart rate), the goodness of fit reached 66%. The same picture was observed using the model derived from of all 59 study-specific data points. No significant relationships were also observed between the changes in slopes estimates and absolute humidity (*p* = 0.26) or the composite score of exercise stress (*p* = 0.65).

**Figure 4.**
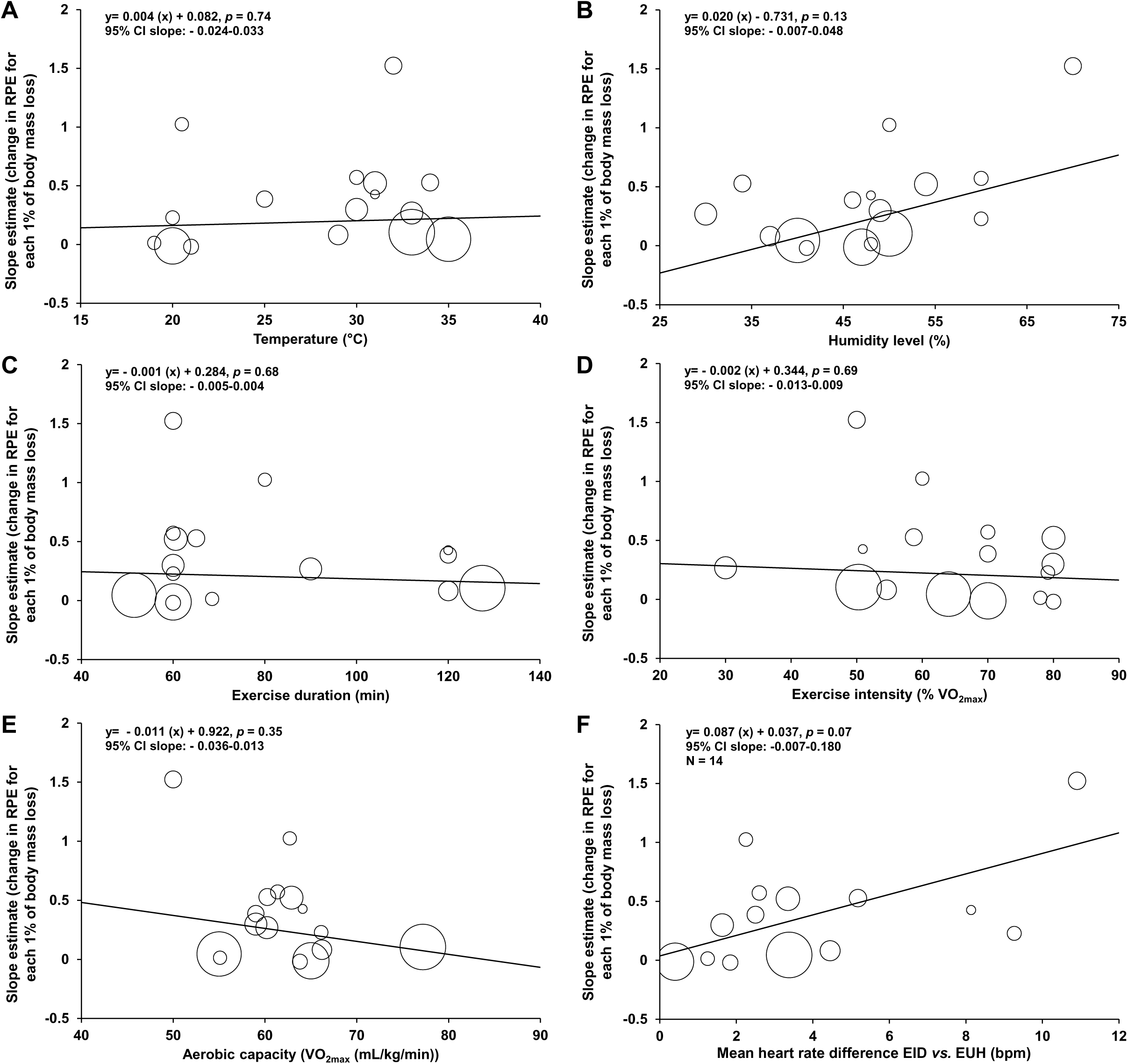
Relationships between the changes in slope estimates and (a) temperature, (b) humidity level, (c) exercise duration, (d) exercise intensity, (e) aerobic capacity (⩒O_2max_) and mean (f) heart rate difference across the different studies included in the meta-analysis. Otherwise stated n = 16.

## DISCUSSION

Despite that exercisers have access to a large and diversified arsenal of tools to monitor exercise intensity, the ability to maintain a certain speed or power output is, ultimately, tributary to RPE (26, 35, 77). On the other hand, enjoyment of exercise is an important component of participation and adherence, and the more the exercise is perceived to be strenuous, the less likely the exercise behaviour is to be maintained (33). In a sense, therefore, RPE can be considered as the ‘’mastermind’’ of exercise performance or adherence. Consequently, factors susceptible to negatively impact the sense of effort during exercise should be given particular attention. Albeit the current results do show that EID increases RPE (response to question #1), the effect was shown to be below our identified threshold of 1 point, at least for the included studies where the greatest level of EID reached was 3% (response to questions #2 and #3), and not moderated by key confounders or associated with changes in heart rate (response to question #4 and #5). Therefore, our results highlight for the first time that EID has a spurious effect on RPE, contrarily to what is believed.

It has been suggested that RPE may act as a mediator of the effect of EID on endurance performance (24, 55). This is legitimate from a physiological perspective, as the documented impact of EID on thermoregulatory, cardiovascular and metabolic functions (24, 55) should result in a higher perceived strain and, hence, RPE. However, our results including observations from cycling and running exercises conducted at clamped and self-paced intensities and under different environmental conditions show that although the change in RPE statistically relates to EID, the magnitude of the effect is unlikely to be practically meaningful until a body mass of at least 3%. Of course, the design of the present meta-analysis precludes from inferencing about the repercussion of the EID- induced increase in RPE on endurance performance. Nevertheless, we are aware of no studies which have been able to establish a decisive relationship between RPE and endurance performance. That being said, if scientists agree upon the simplistic and imperfect model indirectly linking the EID-induced increase in RPE with the decline of endurance performance, the present findings clearly dispute this assertion.

It is proposed that EID may reduce exercise participation/adherence in recreationally active individuals because of its impact upon RPE (36, 82). Based on urinary indices, studies have suggested that 40-50% of recreationally active individuals begin exercise in a light hypohydration state (82, 91) and lose ∼ 0.6% of their body mass while drinking fluid *ad libitum* during freely chosen exercise sessions (82). Furthermore, untrained individuals have lower sweat rate (69, 70) and generally an easy access to water during exercise. Regarding women, they generally have a lower sweat rate than men and drink more (sometimes more than their sweat losses) during exercise (10, 80). Therefore, it is unlikely that those individuals will reach EID levels ≥3% of body mass during typical physical activities and, hence, that the EID-associated increase in RPE should not be a cause for concern. Moreover, studies have reported worsened affective response when participants begin exercise in a low hypohydration state (82) or when dehydration occurs progressively during exercise (8). And it seems that the acute affective response to exercise more than the acute change in RPE relates to long-term adherence to exercise (86, 97).

Although it is agreed upon that the brain is the organ responsible for the regulation of RPE, whether it is centrally derived and largely independent of peripheral afferent signals (21, 64, 67, 90) or results from the integration and interpretation of afferent feedback from the peripheral machinery (5, 50, 78, 88) is debated (see (1, 49, 81, 83) for further details). If the first theory is favoured, then it follows that EID would likely not alter the ability of the brain to produce motor forward commands, termed efference copies or corollary discharges (92). It has been shown that acute EID and the associated hyperosmolality does not alter brain volume (96), potentially highlighting the fact that the cerebral cortex operates close to optimally under dehydrating conditions. On the other hand, interpreting the current results within the context of the second theory would imply that EID provokes minimal homeostasis alterations, at least up to a body mass loss of 3%. This claim is reasonable given that the EID-induced increase in heart rate (∼ 4 beats/min, n = 14) and core temperature (∼ 0.2°C, n = 9) (both results not shown) was marginal compared with EUH.

There was substantial heterogeneity among research findings, as illustrated in Figure 3b. This is unremarkable given that studies used a variety of protocols within which factors known to influence RPE were present. Meta-regressions were conducted to examine the moderating effect of *a priori* selected confounders. They showed that, in isolation, neither humidity, exercise duration, exercise intensity, aerobic capacity nor ambient temperature significantly correlated to the extent of changes in RPE for each 1% of body mass loss. However, while aggregated into one model, those 5 variables explained 66% of the variance observed among the changes in RPE for each 1% of body mass loss. This figure is impressive and depicts the importance of the identified confounders in the overall moderation of the relationship between EID and RPE. Hence, it cannot be excluded that the significance of several influential variables within each of the included studies masked the ability to clearly identify the independent moderating effect of some or all of the confounders.

There was a trend for the change in heart rate to be associated with the change in RPE for each 1% of body mass loss. Such an observation was to be expected because EID is known to exacerbate heart rate (3) and the latter has been reported to be closely related to RPE (50). Indeed, the 6-20 Borg scale was initially developed in healthy individuals to correlate approximately with exercise heart rate. Roughly, our model indicates that for each increase in heart rate of 1 beat/min there should be an increase in RPE of 0.1 points for each 1% of body mass loss. Providing that the mean change in heart rate during exercise was < 10 beats/min between EUH and EID, our observation of a lack of a meaningful effect of EID on RPE makes sense. Nevertheless, it is important to bear in mind that the relationship between heart rate and RPE is only correlational in nature, not causal (50). Indeed, research has shown that it is possible to dissociate the change in heart rate from the change in RPE (e.g., using pharmacological agents) (27, 76). Therefore, our observation of a close relationship between the changes in heart rate and RPE for a given body mass loss should not be taken as a possibility that heart rate could act as a mediator of the relationship between EID and RPE.

Results of this meta-analysis must be interpreted with the following considerations or limitations in mind. The literature search was limited to English and French citations; studies published in different languages may have been missed. Similarly, in the literature of concern, RPE is almost exclusively studied as a secondary outcome. This complicates study identification which may have led us to miss key studies. Validity of RPE measurement depends upon the proper instructions provided to participants (49); no studies reported on whether they dispensed such instructions. Little information is available regarding what represents a meaningful change in RPE; therefore, having used a different threshold may have modified our conclusions. While they would have provided insight into the possible mechanisms linking EID to RPE, data such as thirst sensation, changes in plasma osmolality and volume were not considered as they were reported by too few included studies. Finally, the present results apply to adults, primarily males, up to an EID level of 3% body mass and for exercise up to ∼ 2 h in duration in thermoneutral to warm environments.

In conclusion, while from a statistical point of view, EID > 1% of body mass increases RPE, the present results suggest that its effect is unlikely to be practically meaningful under running or cycling exercise conditions until a body mass loss of at least 3% is reached.

## PERSPECTIVES

Perceived exertion can be considered as the ‘’mastermind’’ of exercise performance or adherence. Exercise-induced dehydration is generally thought to increase RPE. The greater RPE associated with EID may contribute to reducing (1) exercise performance in athletes and (2) exercise participation/adherence in recreationally active individuals. Findings of the present meta-analysis suggest that from a statistical point of view, EID > 1% of body mass increases RPE. However, the effect of EID on RPE is unlikely to be practically meaningful under running or cycling exercise conditions, even at 3% of body mass loss. Thus, our results suggest that concerns about the impact of EID upon RPE and, thus, by extension, the effect of the latter on endurance performance or exercise participation/adherence seem not warranted, at least not until a body mass loss of 3%.

## Data Availability

All data produced in the present study are available upon reasonable request to the authors

## ACKNOWLEDGMENTS

The authors wish to thank the researchers who shared experimental data and provided further information.

No funding was received for the conduct of the work or preparation of the manuscript. TAD is financially supported by the Fonds de Recherche du Québec - Santé (FRQS).

The results of the present study do not constitute endorsement of the product by the authors or the NSCA. The authors have no conflicts of interest to declare.

TAD designed the research, performed the data extraction, performed the statistical analyses, designed the tables and the figures, interpreted data, drafted and revised the manuscript. TP performed the data extraction, interpreted data, drafted and revised the manuscript. EDBG designed the research, performed the data extraction, performed the statistical analyses, designed the tables and figures, interpreted data, drafted and revised the manuscript. All authors have read and approved the final version of the manuscript.

## Keywords and strategy used for the research of potential studies

|  |  |  |
| --- | --- | --- |
| **Hydration (S1)** | Hydrat* or Dehydrat* or Hypohydrat* or Euhydrat* or fluid or drink* | Title or abstract |
| **Perceived exertion (S2)** | “Perceived exertion” or “Perceived effort” or “Rate of perceived exertion” or “Rating of perceived exertion” or “RPE” or percept* or exertion or effort | Title or abstract |
| **Exercise (S3)** | Exercise* or run* or cycl* or endurance or performance | Title or abstract |
| **Overall research** | S1 AND S2 AND S3 |  |

*Note: Research has been done on EBSCO with 4 different databases: SPORTDiscus, CINAHL, AMED and MEDLINE. The same research was also done using PubMed. Last search: December 17, 2020*

